# Risk of subsequent primary cancers among adult cancer survivors in Alberta

**DOI:** 10.1101/2024.02.21.24303162

**Authors:** Matthew T. Warkentin, Winson Y. Cheung, Darren R. Brenner, Dylan E. O’Sullivan

## Abstract

**Background:** Improvements in cancer control have led to a drastic increase in cancer survivors who may be at an elevated risk of developing a subsequent primary cancer (SPC). In this study, we assessed the risk and patterns of SPC development among 134,693 adult cancer survivors in Alberta, Canada.

**Methods:** We used data from the Alberta Cancer Registry to identify all first primary cancers (FPC) occurring between 2004 and 2015. A SPC was considered as the next primary cancer occurring in a different site. We estimated standardized incidence ratios (SIR) for SPC development as the observed number of SPC (O) divided by the expected number of SPC (E), where E is a weighted-sum of the population-based year-age-sex-specific incidence rates and the corresponding person-years of follow-up.

**Results:** The risk of developing a SPC up to fifteen years after an initial cancer was 16.1% for males and 12.3% for females, though these estimates vary considerably by cancer site. Survivors of initial head and neck cancers had a 21.3% fifteen-year cumulative incidence and a 2.5-fold relative risk of SPC development. Overall, both males (SIR=1.50) and females (SIR=1.64) had an increased risk of a SPC. There were significant increases in SPC risk for nearly all age groups, with a greater than 5-fold increase for survivors of cancers diagnosed between ages 18-39.

**Conclusions:** Cancer survivors of nearly every FPC site had substantially increased risk of a SPC, compared to the cancer risk in the general population. Screen-detectable cancers (breast, cervical, colorectal, lung) were common SPC sites and highlights the need to investigate optimal strategies for screening the growing population of cancer survivors.

## Introduction

With a growing and aging Canadian population, the total number of incident cancers has been steadily increasing for many years. There is expected to be 239,100 new cancers in Canada in 2023 alone [1]. In addition, cancer is the leading cause of death in Canada, where one in four Canadians will die from cancer, with an estimated 86,700 cancer deaths expected in 2023 [1]. Despite this poor outlook, survival has been increasing for most cancers for several decades. Advancements in cancer screening, early detection, and treatments, especially in the metastatic setting, have led to significant improvements in survival for many cancers. This trend is expected to continue in the future with ongoing advancements in cancer control.

It is estimated that 1.5 million Canadians are currently living with and beyond cancer [2]. However, cancer survivors are at an elevated risk of developing a new or second primary cancer (SPC) during their lifetime compared to the risk of developing a first cancer among the general population [3]. The heightened risk of developing a subsequent primary cancer may be due, in part, to common exposures across cancers (e.g., smoking), shared genetics, or sequela from cancer treatments (e.g., chemotherapy and radiation) [4]. Despite carrying a higher risk of developing a new cancer, screening guidelines for cancer survivors are largely the same as those for the general population, and adherence to screening among survivors is suboptimal [5]. Moreover, while cancer survivors may undergo surveillance that targets the anatomical site of the initial primary cancer, it is not standard practice for surveillance to be increased for other sites.

In Canada, it is not currently known the rate at which subsequent cancers occur among adult cancer survivors and the relative risk when compared to that observed in the general population. Several studies outside Canada have investigated the risk of SPC development [6–10], however, to our knowledge, there are no Canadian data comprehensively characterizing the landscape of SPC risk. The goal of this study was to estimate the risk of developing a SPC among adult cancer survivors in Alberta, Canada, with an emphasis on screen-detectable cancers. Our findings may inform future screening guidelines and estimate the impact of SPC on the healthcare system.

## Methods

### Data Sources

Data for this study were obtained from the Alberta Cancer Registry (ACR). The ACR is a population-based cancer registry that covers all cancers for the province of Alberta, Canada. ACR has received gold certification from the North American Association of Central Cancer Registries (NAACCR) since 2005. We included all cancer diagnosis occurring between 2004 and 2020 in those aged 18 years or older at the time of their first cancer diagnosis. First primary cancers (FPC) had to occur on December 31st, 2015 or earlier to allow for an adequate follow-up to assess subsequent primary cancers (SPC) among the most recent FPC diagnoses. FPC patients were required to survive at least six months (*i.e.*, minimum of 180 days) from their diagnosis date to be eligible for inclusion in this study. FPC patients who were still alive at the end of follow-up and had not yet been diagnosed with an SPC were right-censored on December 31st, 2020 as the data used in this study had no cancer information beyond this date and a SPC diagnosis could not be ruled out in this period. We excluded all stage 0 cancers (*in situ*) except for bladder cancers. Age at cancer diagnosis was recoded into five-year age bands except for the 18-24 and 90+ age groups. For reporting, cancer sites were recoded to align with the definitions reported by the Canadian Cancer Statistics (CCS) reports (See **Supplemental Table 1**). Alberta population counts stratified by year, age, and sex were obtained from the Statistics Canada Web Data Service API to estimate population-level cancer incidence rates. Ethical approval for this study was obtained from the Health Research Ethics Board of Alberta (HREBA) - Cancer Committee (HREBA.CC-23-0133).

### Statistical Analysis

General population incidence rates were estimated based on the year-age-sex-specific number of cancers in the ACR divided by the corresponding population counts. For any given cancer site, general population incidence rates were estimated excluding that cancer site to estimate risk of developing any cancer other than that site. Metachronous different-site SPC were considered as any SPC occurring at a different cancer site (using the more granular ACR cancer site definitions) more than 180 days after the diagnosis date of the FPC. Follow-up time was defined as 180 days from the date of the FPC until the diagnosis date of the SPC, death, or the end of follow-up (December 31st, 2020), whichever occurred first. Screen-detectable cancers included colorectal, lung, cervix, and breast cancers. Cumulative incidence functions were estimated for risk of SPC or the competing risk of death from any cause by sex and FPC site.

Patient characteristics for cancer survivors are reported as means and standard deviations for continuous variables and frequencies and proportions for categorical variables, separately by sex. The observed number of SPC were the counts of the number of SPC among FPC survivors. The expected number of SPC were estimated as a weighted-sum of the year-age-sex-specific general population incidence rates multiplied by the amount of follow-up time observed among FPC survivors. Where applicable, all results were estimated and reported separately by sex. Incidence rates (per 10,000 person-years) were estimated as the observed number of SPC divided by the total amount of person-years follow-up, multiplied by 10,000. Confidence intervals for incidence rates were obtained using Poisson regression. Standardized incidence ratios (SIR) were estimated as the ratio of the observed number of cancers to the expected number of cancers. Confidence intervals for SIR were estimated using a normal approximation. Survivors were grouped into quartiles based on neighborhood-level annual household income. We assessed whether risk of SPC differed by income quartiles or location of residence (urban vs. rural). All statistical analyses were performed using R version 4.2.1 (2022-06-23).

## Results

In this study, there were 134,693 cancer survivors, 63,340 females and 71,353 males, with a total of 852,661 years of person-time follow-up (average of 6.3 years of follow-up per survivor). Descriptive statistics for the cohort of cancer survivors included in this study are presented in **Table 1**. Among these individuals, there were 13,282 SPC diagnosed during follow-up. **Table 2** presents the number of cancer survivors, average age at FPC and SPC, number of observed and expected SPC, as well as the incidence rate and standardized incidence ratios with corresponding 95% CI, by sex and cancer site. There was an increased relative risk of a SPC overall (SIR=1.55, 95% CI: 1.53-1.58), and for both males (SIR=1.50, 95% CI: 1.47-1.53) and females (SIR=1.64, 95% CI: 1.60-1.69). For females, the largest number of cancer survivors were breast (n=22,690), colorectal (n=6,820), and lung (n=5,955) which had SIR for relative risk of a SPC of 1.46 (95% CI: 1.42-1.57), 1.44 (95% CI: 1.33-1.56), and 1.46 (95% CI: 1.28-1.64), respectively. For males, the largest number of cancer survivors was for prostate (n=24,940), colorectal (n=8,897), and lung (n=5,333), with SIR of 1.31 (95% CI: 1.26-1.35), 1.21 (95% CI: 1.13-1.29), and 1.20 (95% CI: 1.06-1.36), respectively. The largest site-specific SIR was head and neck FPC among females (SIR=2.73, 95% CI: 2.33-3.17) and males (SIR=2.45, 95% CI: 2.23-2.69).

**Table 1.** Descriptive statistics for adult cancer survivors in Alberta diagnosed between 2004-2015, separately by sex. Means and standard deviations are reported for continuous variables, frequencies and proportions are reported for categorical variables.

|  | **Overall**<br>**(N = 134,693)** | **Females**<br>**(N = 63,340)** | **Males**<br>**(N = 71,353)** |
| --- | --- | --- | --- |
| Person-years follow-up<sup>a</sup> | 6.3 [4.5] | 6.5 [4.5] | 6.2 [4.5] |
| Age at FPC diagnosis | 63.4 [13.9] | 62.4 [14.7] | 64.3 [13.1] |
| TNM Stage for FPC |  |  |  |
| I | 32,636 (24.2%) | 21,022 (33.2%) | 11,614 (16.3%) |
| II | 32,134 (23.9%) | 12,789 (20.2%) | 19,345 (27.1%) |
| III | 18,572 (13.8%) | 9,444 (14.9%) | 9,128 (12.8%) |
| IV | 16,309 (12.1%) | 6,876 (10.9%) | 9,433 (13.2%) |
| Other<sup>b</sup> | 35,042 (26.0%) | 13,209 (20.9%) | 21,833 (30.6%) |
| Year of diagnosis for FPC |  |  |  |
| 2004-2009 | 62,445 (46.4%) | 29,079 (45.9%) | 33,366 (46.8%) |
| 2010-2015 | 72,248 (53.6%) | 34,261 (54.1%) | 37,987 (53.2%) |
| Cancer site for FPC |  |  |  |
| Bladder | 6,763 (5.0%) | 1,590 (2.5%) | 5,173 (7.2%) |
| Brain/CNS | 1,578 (1.2%) | 626 (1.0%) | 952 (1.3%) |
| Breast | 22,816 (16.9%) | 22,690 (35.8%) | 126 (0.2%) |
| Cervix | 1,446 (1.1%) | 1,446 (2.3%) | — |
| Colorectal | 15,717 (11.7%) | 6,820 (10.8%) | 8,897 (12.5%) |
| Esophagus | 1,062 (0.8%) | 213 (0.3%) | 849 (1.2%) |
| Head and neck | 4,113 (3.1%) | 1,160 (1.8%) | 2,953 (4.1%) |
| Hodgkin lymphoma | 872 (0.6%) | 353 (0.6%) | 519 (0.7%) |
| Kidney and renal pelvis | 4,342 (3.2%) | 1,626 (2.6%) | 2,716 (3.8%) |
| Leukemia | 4,404 (3.3%) | 1,743 (2.8%) | 2,661 (3.7%) |
| Liver | 1,195 (0.9%) | 295 (0.5%) | 900 (1.3%) |
| Lung and bronchus | 11,288 (8.4%) | 5,955 (9.4%) | 5,333 (7.5%) |
| Melanoma | 6,268 (4.7%) | 3,074 (4.9%) | 3,194 (4.5%) |
| Multiple myeloma | 1,694 (1.3%) | 696 (1.1%) | 998 (1.4%) |
| Non-Hodgkin lymphoma | 5,712 (4.2%) | 2,530 (4.0%) | 3,182 (4.5%) |
| Ovary | 1,608 (1.2%) | 1,608 (2.5%) | — |
| Pancreas | 1,700 (1.3%) | 879 (1.4%) | 821 (1.2%) |
| Prostate | 24,940 (18.5%) | — | 24,940 (35.0%) |
| Stomach | 1,655 (1.2%) | 560 (0.9%) | 1,095 (1.5%) |
| Testis | 1,337 (1.0%) | — | 1,337 (1.9%) |
| Uterus | 4,776 (3.5%) | 4,776 (7.5%) | — |
| All other cancers | 9,407 (7.0%) | 4,700 (7.4%) | 4,707 (6.6%) |

Abbreviations: CNS, central nervous system; TNM, tumor-node-metastases classification.
<sup>a</sup> Person-year follow-up was defined as six months from the FPC diagnosis date until date of SPC diagnosis, death, last known date of contact, or December 31<sup>st</sup>, 2020, whichever occurred first.
<sup>b</sup> Other TNM stage includes cancers with stage 0, occult, unknown, or missing.

**Table 2.** The risk of developing a different-site subsequent primary cancer (SPC) among adult cancer survivors in the Alberta Cancer <u>Registry (ACR) diagnosed between 2004-2015.</u>.

| FPC Site | Survivors | Follow-up (PY) | Mean age at FPC | Observed SPC | Mean age at SPC | IR (95% CI) | Expected SPC | SIR (95% CI) |
| --- | --- | --- | --- | --- | --- | --- | --- | --- |
| **OVERALL** | 134,693 | 852,661 | 63.4 | 13,282 | 71.1 | 156 (153-158) | 8,552 | 1.55 (1.53-1.58) |
| **FEMALES** |  |  |  |  |  |  |  |  |
| **Overall** | 63,340 | 411,726 | 62.4 | 5,211 | 69.9 | 127 (123-130) | 3,175 | 1.64 (1.60-1.69) |
| Bladder | 1,590 | 10,415 | 69.0 | 196 | 75.0 | 188 (163-216) | 122 | 1.61 (1.39-1.84) |
| Brain/CNS | 626 | 2,603 | 51.1 | 28 | 60.0 | 108 (72-152) | 11 | 2.64 (1.75-3.71) |
| Breast | 22,690 | 174,300 | 60.5 | 1,623 | 69.0 | 93 (89-98) | 1,088 | 1.49 (1.42-1.57) |
| Cervix | 1,446 | 10,802 | 47.1 | 82 | 57.4 | 76 (61-94) | 41 | 2.00 (1.59-2.45) |
| Colorectal | 6,820 | 40,717 | 67.3 | 597 | 73.5 | 147 (135-159) | 413 | 1.44 (1.33-1.56) |
| Esophagus | 213 | 547 | 66.9 | 14 | 68.6 | 256 (144-414) | 5 | 2.59 (1.41-4.13) |
| Head and neck | 1,160 | 6,847 | 62.2 | 165 | 70.2 | 241 (206-280) | 60 | 2.73 (2.33-3.17) |
| Hodgkin lymphoma | 353 | 3,067 | 40.7 | 24 | 60.7 | 78 (51-114) | 10 | 2.40 (1.53-3.45) |
| Kidney and renal pelvis | 1,626 | 11,083 | 62.2 | 190 | 69.3 | 171 (148-197) | 98 | 1.93 (1.67-2.21) |
| Leukemia | 1,743 | 12,109 | 63.2 | 218 | 71.2 | 180 (157-205) | 119 | 1.84 (1.60-2.09) |
| Liver | 295 | 959 | 66.5 | 13 | 67.3 | 136 (75-223) | 10 | 1.30 (0.69-2.11) |
| Lung and bronchus | 5,955 | 17,606 | 67.9 | 255 | 71.6 | 145 (128-163) | 175 | 1.46 (1.28-1.64) |
| Melanoma | 3,074 | 25,858 | 56.4 | 260 | 66.1 | 101 (89-113) | 179 | 1.45 (1.28-1.64) |
| Multiple myeloma | 696 | 3,236 | 68.1 | 58 | 70.2 | 179 (137-229) | 36 | 1.59 (1.21-2.03) |
| Non-Hodgkin lymphoma | 2,530 | 16,887 | 63.8 | 251 | 69.5 | 149 (131-168) | 160 | 1.57 (1.38-1.77) |
| Ovary | 1,608 | 7,596 | 58.7 | 52 | 63.8 | 68 (51-89) | 54 | 0.97 (0.72-1.25) |
| Pancreas | 879 | 1,576 | 66.9 | 22 | 63.9 | 140 (89-206) | 16 | 1.34 (0.84-1.96) |
| Stomach | 560 | 2,077 | 66.0 | 41 | 71.7 | 197 (143-264) | 21 | 1.95 (1.40-2.59) |
| Uterus | 4,776 | 35,948 | 61.6 | 441 | 69.4 | 123 (112-134) | 305 | 1.44 (1.31-1.58) |
| All other cancers | 4,700 | 24,490 | 64.8 | 681 | 71.7 | 248 (230-267) | 251 | 2.71 (2.51-2.92) |
| **MALES** |  |  |  |  |  |  |  |  |
| **Overall** | 71,353 | 440,935 | 64.3 | 8,071 | 72.0 | 183 (179-187) | 5,376 | 1.50 (1.47-1.53) |
| Bladder | 5,173 | 5,173 | 68.9 | 924 | 74.5 | 301 (282-321) | 516 | 1.79 (1.68-1.91) |
| Brain/CNS | 952 | 3,459 | 51.0 | 22 | 59.0 | 64 (41-94) | 14 | 1.57 (0.98-2.29) |
| Breast | 126 | 753 | 66.9 | 17 | 75.2 | 226 (135-351) | 12 | 1.41 (0.82-2.16) |
| Colorectal | 8,897 | 51,177 | 65.6 | 885 | 73.1 | 173 (162-185) | 734 | 1.21 (1.13-1.29) |
| Esophagus | 849 | 1,816 | 64.5 | 41 | 67.6 | 226 (164-302) | 25 | 1.63 (1.17-2.16) |
| Head and neck | 2,953 | 16,360 | 60.5 | 443 | 68.7 | 271 (246-297) | 181 | 2.45 (2.23-2.69) |
| Hodgkin lymphoma | 519 | 4,317 | 39.8 | 30 | 55.5 | 69 (47-97) | 13 | 2.28 (1.54-3.17) |
| Kidney and renal pelvis | 2,716 | 17,034 | 61.0 | 346 | 69.1 | 203 (182-225) | 199 | 1.74 (1.56-1.93) |
| Leukemia | 2,661 | 16,896 | 62.7 | 395 | 70.1 | 234 (211-258) | 227 | 1.74 (1.57-1.92) |
| Liver | 900 | 3,057 | 62.0 | 42 | 64.2 | 137 (100-183) | 37 | 1.13 (0.82-1.50) |
| Lung and bronchus | 5,333 | 12,512 | 68.7 | 243 | 73.7 | 194 (171-220) | 202 | 1.20 (1.06-1.36) |
| Melanoma | 3,194 | 23,596 | 60.6 | 377 | 71.4 | 160 (144-176) | 276 | 1.37 (1.23-1.51) |
| Multiple myeloma | 998 | 4,805 | 65.9 | 86 | 70.7 | 179 (144-219) | 71 | 1.21 (0.97-1.48) |
| Non-Hodgkin lymphoma | 3,182 | 20,408 | 61.0 | 371 | 69.3 | 182 (164-201) | 246 | 1.51 (1.36-1.67) |
| Pancreas | 821 | 1,499 | 64.6 | 24 | 68.6 | 160 (104-233) | 21 | 1.15 (0.73-1.65) |
| Prostate | 24,940 | 193,154 | 66.1 | 2,845 | 73.1 | 147 (142-153) | 2,179 | 1.31 (1.26-1.35) |
| Stomach | 1,095 | 3,573 | 66.2 | 68 | 73.7 | 190 (149-239) | 60 | 1.14 (0.88-1.43) |
| Testis | 1,337 | 11,924 | 34.7 | 34 | 47.6 | 29 (20-39) | 15 | 2.21 (1.53-3.02) |
| All other cancers | 4,707 | 23,939 | 66.1 | 878 | 72.2 | 367 (343-392) | 349 | 2.52 (2.35-2.68) |

⇒ Abbreviations: CI, confidence interval; CNS, central nervous system; FPC, first primary cancer; IR, incidence rate; NOS, not otherwise specified; PY, person-SIR, standardized incidence ratio; SPC, subsequent primary cancer.

The cumulative incidence for developing a SPC by sex while accounting for the competing risk of all-cause mortality is presented in **Figure 1**. The risk of a SPC is 6.9%, 12.1%, and 16.1% at 5, 10, and 15-years since the FPC diagnosis for males, respectively (see **Figure 1A**). The risk of a SPC is 4.8%, 8.7%, and 12.3% at 5, 10, and 15-years since the FPC diagnosis for females, respectively. The cumulative incidences for a SPC by sex at several time horizons based on the location of the FPC are presented in **Figure 1B**.

**Figure 1.**
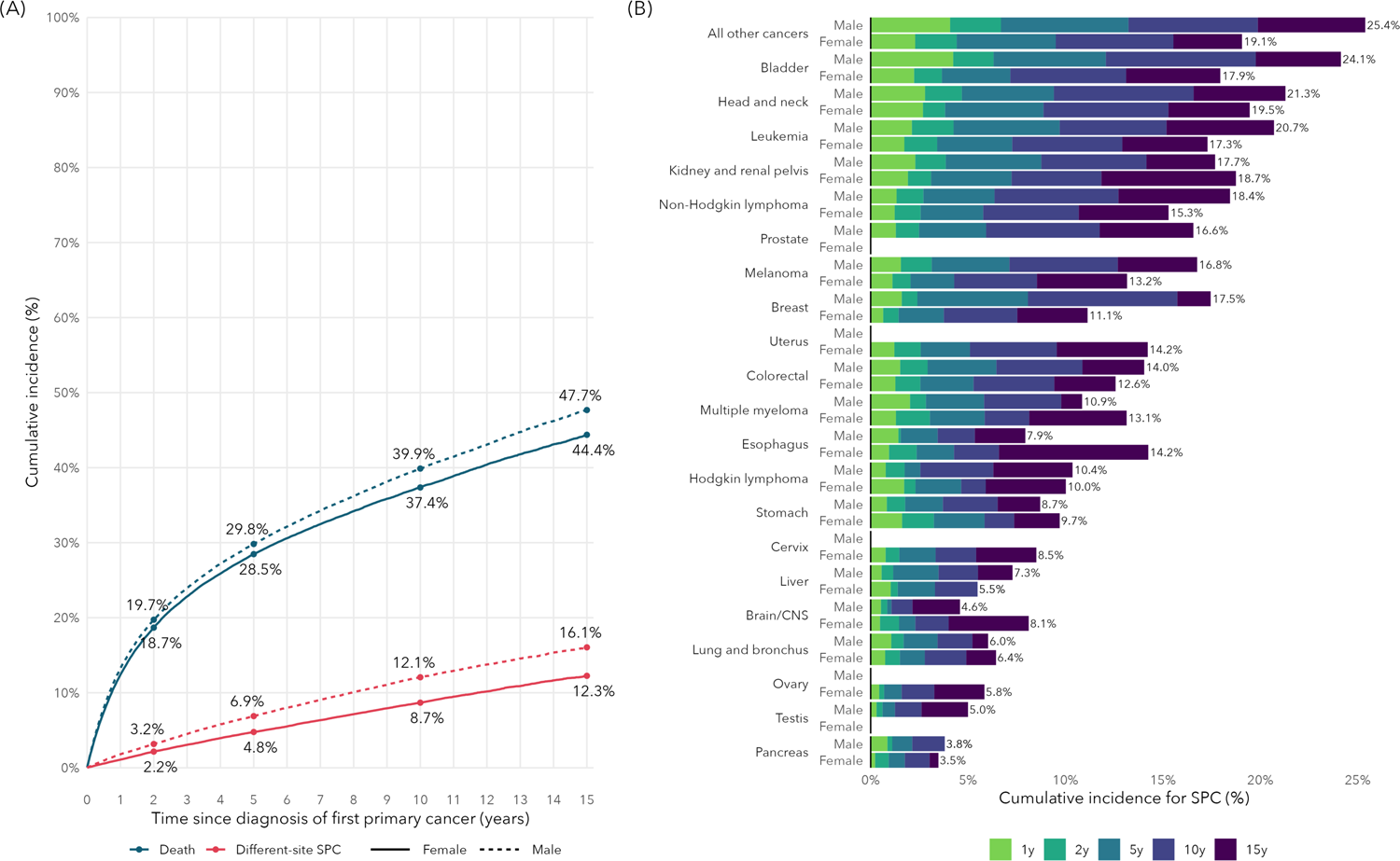
**(A)** Cumulative incidence for death (from any cause) or the development of a different-site second primary cancer (SPC) for cancer survivors in Alberta, separately by sex. **(B)** Cumulative incidence for a different-site SPC at 1, 2, 5, 10, and 15-year time horizons based on the location of the first primary cancer (FPC), separately by sex.

We present the SIR for risk of SPC by age at diagnosis for the FPC by sex in **Figure 2** and **Supplemental Table 2**. There was a trend of higher relative risk among those diagnoses with FPC at an early age, and the risk attenuated for older age groups. Similarly, we present the relative risk for a SPC by year of diagnosis for the FPC, separately by sex, in **Table 3**. In general, the number of cancer survivors increased over time (see **Supplemental Figure 1**), while the relative risk remained relatively stable. Lastly, we present the relative risk for a site-specific SPC for any FPC by sex in **Table 4**. For females, multiple myeloma, leukemia, and esophagus cancer had more than a 2-fold relative risk after a FPC. These findings were similar for males, however, Hodgkin lymphoma carried two-fold relative risk while esophageal cancer did not. However, the most common SPC sites were lung (n=931, 17.9%), breast (n=845, 16.2%), and colorectal (n=592, 11.4%) for females, and prostate (n=1,244, 15.4%), lung (n=1,233, 15.3%), and colorectal (991, 12.3%) for males. We present risk of SPC stratified by geographic location and income quartiles in **Supplemental Table 3**; the relative risks (SIR) were fairly constant across these socioeconomic strata.

**Figure 2.**
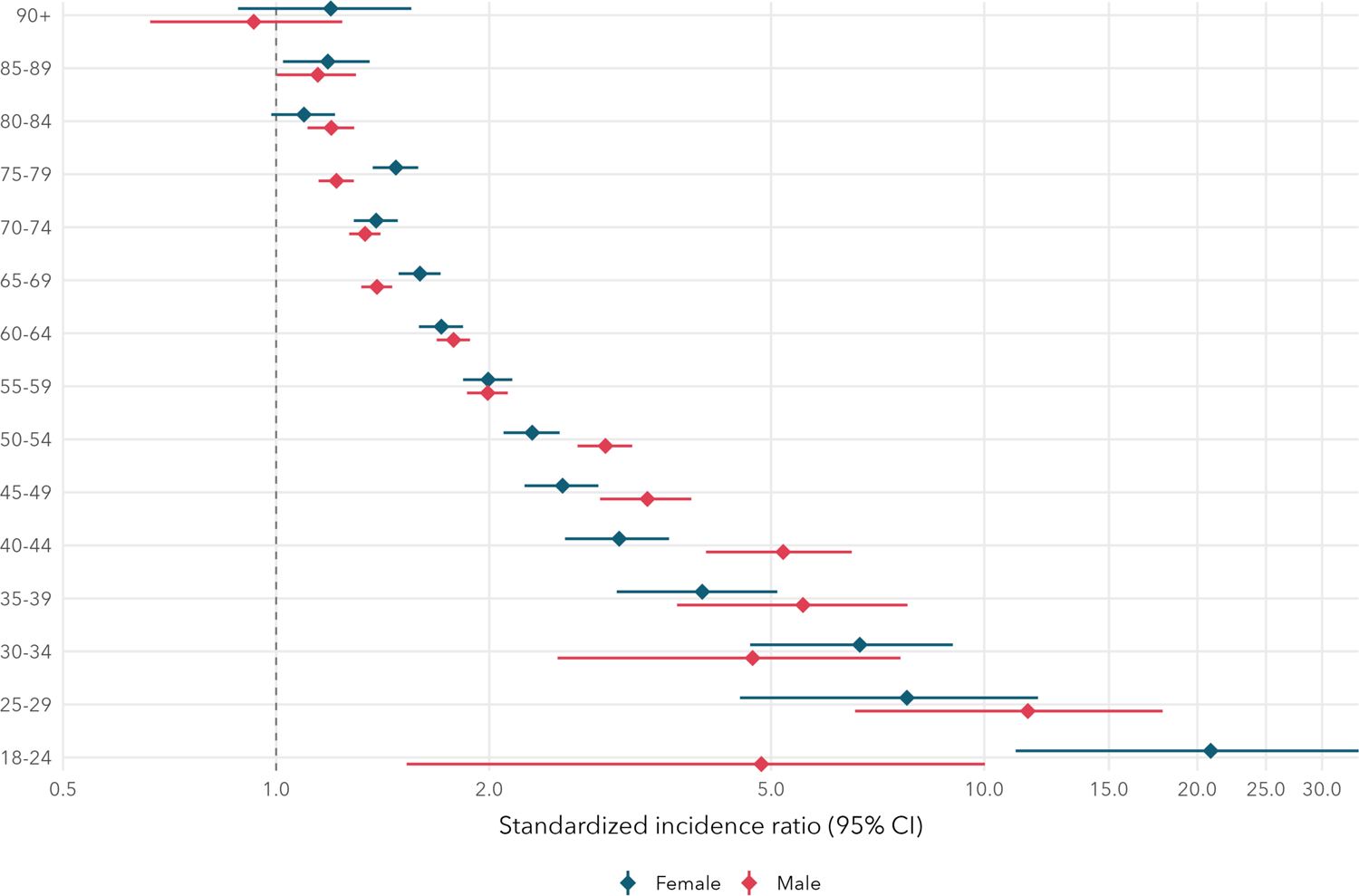
Standardized incidence ratios (SIR) and 95% confidence intervals for the risk of a different-site subsequent primary cancer (SPC) based on the age at diagnosis for the first-primary cancer (FPC), separately by sex.

**Table 3.** Standardized incidence ratios (SIR) for risk of a different-site subsequent <u>primary cancer by year of first primary cancer diagnosis and sex.</u>.

|  | Females |  | Males |  |
| --- | --- | --- | --- | --- |
|  | Survivors | SIR (95% CI) | Survivors | SIR (95% CI) |
| **2004** | 4,758 | 1.75 (1.61-1.91) | 5,338 | 1.52 (1.41-1.62) |
| **2005** | 4,741 | 1.62 (1.48-1.77) | 5,130 | 1.59 (1.48-1.70) |
| **2006** | 4,800 | 1.77 (1.62-1.92) | 5,540 | 1.45 (1.35-1.55) |
| **2007** | 4,881 | 1.80 (1.65-1.97) | 5,612 | 1.53 (1.42-1.64) |
| **2008** | 4,841 | 1.67 (1.52-1.83) | 5,822 | 1.43 (1.33-1.54) |
| **2009** | 5,058 | 1.75 (1.60-1.92) | 5,924 | 1.54 (1.44-1.66) |
| **2010** | 5,256 | 1.66 (1.51-1.82) | 5,883 | 1.63 (1.51-1.75) |
| **2011** | 5,376 | 1.51 (1.37-1.67) | 6,285 | 1.42 (1.31-1.53) |
| **2012** | 5,429 | 1.66 (1.50-1.82) | 6,327 | 1.47 (1.36-1.59) |
| **2013** | 5,668 | 1.58 (1.43-1.75) | 6,174 | 1.54 (1.42-1.67) |
| **2014** | 6,245 | 1.36 (1.22-1.51) | 6,622 | 1.37 (1.25-1.49) |
| **2015** | 6,287 | 1.44 (1.28-1.61) | 6,696 | 1.52 (1.39-1.66) |

Abbreviations: CI, confidence interval; SIR, standardized incidence ratio

**Table 4.** Standardized incidence ratios (SIR) for risk of a site-specific subsequent primary cancer by sex, based on any different-site FPC.

|  | Females |  |  | Males |  |  |
| --- | --- | --- | --- | --- | --- | --- |
|  | Different-site survivors | O/E | SIR (95% CI) | Different-site survivors | O/E | SIR (95% CI) |
| **Bladder** | 61,750 | 177/100 | 1.78 (1.52-2.05) | 66,180 | 681/448 | 1.52 (1.41-1.64) |
| **Brain/CNS** | 62,714 | 57/40 | 1.44 (1.09-1.83) | 70,401 | 85/75 | 1.14 (0.91-1.39) |
| **Breast** | 40,650 | 844/641 | 1.32 (1.23-1.41) | 71,227 | 20/10 | 1.91 (1.17-2.84) |
| **Cervix** | 61,894 | 73/44 | 1.66 (1.30-2.06) | — | — | — |
| **Colorectal** | 56,520 | 556/387 | 1.44 (1.32-1.56) | 62,456 | 950/730 | 1.30 (1.22-1.38) |
| **Esophagus** | 63,127 | 47/21 | 2.28 (1.68-2.98) | 70,504 | 169/105 | 1.61 (1.37-1.86) |
| **Head and neck** | 62,180 | 104/67 | 1.56 (1.27-1.87) | 68,400 | 274/217 | 1.26 (1.12-1.42) |
| **Hodgkin lymphoma** | 62,987 | 15/8 | 1.78 (0.99-2.79) | 70,834 | 27/14 | 2.00 (1.32-2.82) |
| **Kidney and renal pelvis** | 61,714 | 112/95 | 1.18 (0.97-1.41) | 68,637 | 321/219 | 1.47 (1.31-1.63) |
| **Leukemia** | 61,597 | 286/108 | 2.64 (2.34-2.95) | 68,692 | 477/227 | 2.10 (1.92-2.29) |
| **Liver** | 63,045 | 45/27 | 1.67 (1.22-2.19) | 70,453 | 121/102 | 1.18 (0.98-1.40) |
| **Lung and bronchus** | 57,385 | 931/561 | 1.66 (1.56-1.77) | 66,020 | 1,233/887 | 1.39 (1.31-1.47) |
| **Melanoma** | 60,266 | 177/125 | 1.42 (1.21-1.63) | 68,159 | 343/217 | 1.58 (1.42-1.75) |
| **Multiple myeloma** | 62,644 | 155/51 | 3.02 (2.57-3.52) | 70,355 | 247/99 | 2.50 (2.20-2.82) |
| **Non-Hodgkin lymphoma** | 60,810 | 280/148 | 1.89 (1.67-2.12) | 68,171 | 466/254 | 1.83 (1.67-2.01) |
| **Ovary** | 61,732 | 105/93 | 1.12 (0.92-1.35) | — | — | — |
| **Pancreas** | 62,461 | 197/118 | 1.67 (1.45-1.91) | 70,532 | 250/158 | 1.58 (1.39-1.78) |
| **Prostate** | — | — | — | 46,413 | 1,244/1,036 | 1.20 (1.14-1.27) |
| **Stomach** | 62,780 | 78/49 | 1.61 (1.27-1.98) | 70,258 | 208/140 | 1.48 (1.29-1.69) |
| **Testis** | — | — | — | 70,016 | 16/12 | 1.36 (0.77-2.11) |
| **Uterus** | 58,564 | 371/223 | 1.67 (1.50-1.84) | — | — | — |
| **All other cancers** | 58,640 | 459/270 | 1.70 (1.55-1.86) | 66,646 | 732/425 | 1.72 (1.60-1.85) |

Abbreviations: CI, confidence interval; CNS, central nervous system; E, expected number of SPC; O, observed number of SPC; SIR, standardized incidence ratios.

Among the 5,211 and 8,071 SPC among females and males, respectively, 47% of female and 28% of male SPC were among these potentially screen-detectable cancers. Among the 118,976 non-colorectal FPC, there were 1,506 colorectal SPC compared to the 1,117 expected CRC (Female SIR=1.44; Male SIR=1.30). Among the 123,405 non-lung FPC, there were 2,164 lung FPC compared to the 1,448 expected lung cancers (Female SIR=1.66; Male SIR=1.39). Among the 61,894 non-cervical FPC, there were 73 cervical FPC compared to the 44 expected cervical cancers (Female SIR=1.66). Among the 40,650 female non-breast FPC, there were 844 female breast FPC compared to the 641 expected female breast cancers (Female SIR=1.32).

## Discussion

In this study, we leveraged the high-quality and comprehensive province-wide cancer registry data to describe the risk of developing a different-site SPC among cancer survivors in Alberta, Canada. We found that cancer survivors of nearly all FPC sites carry an increased risk of a SPC compared to the general population risk. To contextualize our findings, we will compare our results with those from other international studies, contextualize our findings, describe important gaps in the existing literature for SPC, and the growing importance of surveillance of cancer survivors.

Age is considered one of the strongest predictors for cancer development, especially for common cancers (*e.g.*, lung, breast, colorectal, prostate). In this study, we observed a strong age effect whereby individuals with a first cancer diagnosis at a younger age carried a much higher relative risk of developing a subsequent cancer compared to the general population at the same age. This effect was attenuated in older groups whereby survivors aged 90+ did not exhibit a higher risk of developing a cancer than the general population. This observation may be due to the long latency period for second cancers and late treatment effects to emerge. Standard follow-up for most FPC is 5 years even though young adult patients continue to carry substantial risk of a SPC well beyond this time frame. A US study of young adults who survived at least 5 years observed that breast, lung, and colorectal cancer were 3 of the 4 most common SPC among this patient population and that young adults carried a higher risk of developing late-stage breast and lung cancer than the general population [11]. Future research that focuses on risk factors for screen-detectable SPC are required to determine if risk-stratification approaches would be feasible among cancer survivors.

In general, women had a higher relative risk of developing a SPC (compared to the general population of women) than men (compared to the general population of men), but men were observed to have a higher cumulative lifetime (*i.e.*, absolute) risk of developing a SPC. This may be partly related to the longer life expectancy of women which provide more time and opportunity for the onset of a second primary cancer, as well as the general population of men having a higher cancer risk than the general population of women. For sex-specific cancers, female cancers of the breast, cervix, and uterus all carried significantly increased risk compared to the general population. For men, an initial testicular cancer carried a more than 2-fold increase in risk for an SPC, while prostate cancer only carried a moderately higher risk than the general population (SIR=1.31). High mortality cancers including ovarian, pancreas, and liver did not carry any increased risk compared to the general population. There was an increased risk of a SPC for all neighborhood income quartiles with no significant change in relative risk across income groups; these findings were consistent with the analyses comparing risk in those dwelling in urban versus rural residences. While the overall risk of developing a SPC is consistent among these groups, the risk may vary by FPC-SPC site combinations, stage of SPC, and mortality associated with SPC, which should be explored in future studies.

One noteworthy finding in our study was the substantial increase in risk for developing a SPC among blood-related cancers (Hodgkin and Non-Hodgkin lymphoma, leukemia, multiple myeloma). In particular, Hodgkin lymphoma survivors carried a more than 2-fold risk of SPC in both men and women. The average age of FPC diagnosis for Hodgkin lymphoma was 40.7 and 39.8 for women and men, respectively. The late and long-term effects of chemotherapy that are often given for extended periods of time for hematologic malignancies may be a contributor to the increased risk of SPC for survivors of these cancers. Additionally, blood cancers were also common among survivors of solid cancers. Cancer survivors who develop subsequent blood cancers are likely a very complex patient population to manage clinically. Future studies should examine the treatment patterns and outcomes of this unique patient population.

There have been several studies performed internationally investigating the risk SPC development. A study in France included 289,967 cancer survivors and observed 21,226 SPC [6]. The relative risk of a SPC was 1.36 (95% CI: 1.35-1.38). They attributed much of the increased risk of a SPC to the high rate of tobacco and alcohol consumption in France [6]. They similarly observed an increase in risk for individuals with a younger age at first cancer diagnosis. An Italian study of 165,179 FPC and 11,348 SPC found substantial sex-differences in the locations of the SPC [9]. This study concluded, among other things, that the development of cancer screening and surveillance plans should be considered based on the risk of common SPC sites [9]. One of the largest studies was performed in the United States using the SEER data [8]. This study included more than 1.5 million cancer survivors who survived at least 5 years and observed 156,442 incident SPC and 88,818 SPC deaths. Overall, they found a 11% and 10% increase in risk and a 12% and 33% increase in mortality for men and women, respectively [8]. They observed an increase in risk of SPC for nearly all cancer sites, which is consistent with the findings of our study. A Danish study of 457,334 survivors found a cumulative incidence (risk) of a SPC of 6.3%, 10.5%, and 13.5% at 5, 10, and 15-years after FPC diagnosis [10], which further support our findings, though they included same-site SPC. A Swiss study of 310,113 cancer survivors observed 33,793 SPC [7]. Similar to our study, they found an increase in relative risk for those diagnosed at younger ages and for those with an initial Hodgkin lymphoma [7]. Multiple studies found that cancers of the head and neck carried significantly higher risk of SPC [7,8,10], which is consistent with a more than 2-fold increased risk observed in our study. This is a particularly high-risk survivorship population that could benefit from enhanced surveillance. Future studies should explore risk-stratified surveillance approaches for high-risk survivorship populations. While only 5% of cancer survivors had a bladder FPC, they had a 15-year risk of developing a SPC of 24.1% for males (highest single site) and 17.9% for females (fourth highest single site).

The exceptional progress in reducing cancer mortality rates across most cancers has led to a growing number of cancer survivors. In Canada, there is an estimated 1.5 million people living with and beyond their cancer diagnosis [2] and this number is projected to increase over time. In our study, we found the number of cancer survivors increased 32% in the 10 years from 2005 to 2015 and are projected to increase by 86% in 2040 [12]. In the US, there are an estimated 18.1 million cancer survivors as of 2022 and this is projected to increase by 22.4% to 22.5 million by the year 2032 [13]. With the number of cancer survivors projected to increase globally, substantial effort and resources will be required to improve our understanding of cancer risk after an initial cancer diagnosis and the cancer control strategies to effectively manage this complex patient population.

This study has several strengths. First, this is one of the largest and most comprehensive studies to date to assess SPC risk in a large cohort of Canadians. Second, this study used high-quality population-based data from the Alberta Cancer Registry that captures over 99% of all cancers in the province of Alberta with detailed epidemiologic and clinical data. However, this study has several limitations worth noting. First, the data used for these analyses did not include thyroid cancers and thus, we are unable to assess thyroid as a FPC site, and our observed counts of SPC may be underestimated, meaning that we do not know the risk associated with thyroid FPC and our SPC risk findings may be partially conservative (*i.e.*, risk of SPC may be even higher among cancer survivors). Second, we did not assess the role of systemic treatments and radiation on SPC risk in this study. However, these data are available and will be explored in future studies. Third, we excluded any same-site SPC to avoid potential misclassification due to recurrence since the discrimination between synchronous and metachronous (*e.g.*, different histology or contralateral side of paired organs) same-site cancers were difficult to assess in administrative data and so our findings underestimate the true risk of SPC.

With the number of cancer survivors increasing in Canada, further research is needed to assess risk of developing a SPC. This study found that most FPC sites carry an increased risk of a different-site SPC compared to cancer risk in the general population, and many of these SPC are considered screen-detectable which offers opportunities to mitigate morbidity and mortality due to SPC. Our findings add to the body of literature that suggest that a different surveillance approach may be needed for cancer survivors, which should consider different risk profiles. Future research should continue to investigate optimal strategies for screening the growing population of cancer survivors.

## Additional Information

**Data Availability Statement:** The data that supports the findings of this study are available from the Alberta Cancer Registry, but restrictions apply to the availability of these data, which were used under license for the current study and so are not publicly available. Data from these analyses are available upon request to researchers with ethics approved projects through the Alberta Cancer Registry.

**Author contributions: MTW:** Conceptualization, Methodology, Formal analysis, Data curation, Writing - Original Draft, Writing - Review & Editing; **WYC:** Writing - Review & Editing; **DRB:** Writing - Review & Editing; **DEO:** Conceptualization, Methodology, Data curation, Writing - Original Draft, Writing - Review & Editing, Supervision, Funding acquisition.

**Ethics:** This study was reviewed by the Health Research Ethics Board of Alberta (HREBA) - Cancer Committee (CC) and was given ethics approval (HREBA.CC-23-0133).

**Funding/Support:** This study did not receive any funding.

**Role of the funder/sponsor:** The funding source had no input into study design; in the collection, analysis and interpretation of data; in the writing of the report; and in the decision to submit the article for publication.

**Conflicts of Interest:** The authors declare no conflicts of interest.

## Data Availability

The data that supports the findings of this study are available from the Alberta Cancer Registry, but restrictions apply to the availability of these data, which were used under license for the current study and so are not publicly available. Data from these analyses are available upon request to researchers with ethics approved projects through the Alberta Cancer Registry.

## References

1. Canadian Cancer Statistics Advisory Council in collaboration with the Canadian Cancer Society, Statistics Canada, and the Public Health Agency of Canada. Canadian cancer statistics 2023. Canadian Cancer Society; 2023.

2. Canadian Cancer Statistics Advisory Council in collaboration with the Canadian Cancer Society, Statistics Canada, and the Public Health Agency of Canada. Canadian cancer statistics: A 2022 special report on cancer prevalence. Canadian Cancer Society; 2022.

3. Murphy CC, Gerber DE, Pruitt SL. Prevalence of prior cancer among persons newly diagnosed with cancer: An initial report from the surveillance, epidemiology, and end results program. JAMA oncology. 2018;4(6):832–6.

4. Wood ME, Vogel V, Ng A, Foxhall L, Goodwin P, Travis LB. Second malignant neoplasms: Assessment and strategies for risk reduction. Journal of clinical oncology. 2012;30(30):3734–45.

5. Uhlig A, Mei J, Baik I, Meyer C, Uhlig J. Screening utilization among cancer survivors: A meta-analysis. Journal of Public Health. 2018;40(1):129–37.

6. Jégu J, Colonna M, Daubisse-Marliac L, Trétarre B, Ganry O, Guizard A-V, Bara S, Troussard X, Bouvier V, Woronoff A-S, others. The effect of patient characteristics on second primary cancer risk in france. BMC cancer. 2014;14(1):1–14.

7. Feller A, Matthes KL, Bordoni A, Bouchardy C, Bulliard J-L, Herrmann C, Konzelmann I, Maspoli M, Mousavi M, Rohrmann S, others. The relative risk of second primary cancers in switzerland: A population-based retrospective cohort study. BMC cancer. 2020;20:1–15.

8. Sung H, Hyun N, Leach CR, Yabroff KR, Jemal A. Association of first primary cancer with risk of subsequent primary cancer among survivors of adult-onset cancers in the united states. Jama. 2020;324(24):2521–35.

9. Ragusa R, Torrisi A, Di Prima AA, Torrisi AA, Ippolito A, Ferrante M, Madeddu A, Guardabasso V. Cancer prevention for survivors: Incidence of second primary cancers and sex differences—a population-based study from an italian cancer registry. International Journal of Environmental Research and Public Health. 2022;19(19):12201.

10. Kjaer TK, Andersen EAW, Ursin G, Larsen SB, Bidstrup PE, Winther JF, Borre M, Johansen C, Dalton SO. Cumulative incidence of second primary cancers in a large nationwide cohort of danish cancer survivors: A population-based retrospective cohort study. The Lancet Oncology. 2023;

11. Sung H, Siegel RL, Hyun N, Miller KD, Yabroff KR, Jemal A. Subsequent primary cancer risk among 5-year survivors of adolescent and young adult cancers. Journal Of The National Cancer Institute. 2022;114(8):1095–108.

12. Brenner DR, Carbonell C, O’Sullivan DE, Ruan Y, Basmadjian RB, Bu V, Farah E, Loewen SK, Bond TR, Estey A, others. Exploring the future of cancer impact in alberta: Projections and trends 2020–2040. Current Oncology. 2023;30(11):9981–95.

13. Miller KD, Nogueira L, Devasia T, Mariotto AB, Yabroff KR, Jemal A, Kramer J, Siegel RL. Cancer treatment and survivorship statistics, 2022. CA: a cancer journal for clinicians. 2022;72(5):409–36.

