## Supplemental Materials for "Risk of subsequent primary cancers among adult cancer survivors in Alberta"

**Supplemental Table 1.** Mapping between the Canadian Cancer Statistics (CCS) and the Alberta Cancer Registry (ACR) cancer site definitions.

| CCS definition (N = 22) | ACR definition (N = 63) |
|---|---|
| Bladder (C67) | Bladder (C67) |
| Brain/CNS (C70-C72) | Brain (C71) |
| | Spinal Cord, Cranial Nerves, and other CNS (C72) |
| Breast (C50) | Breast (C50) |
| Cervix (C53) | Cervix Uteri (C53) |
| Colorectal (C18-C20, C26.0) | Colorectal (C18, C20) |
| | Rectosigmoid Junction (C19) |
| | Small Intestine (C17) |
| Esophagus (C15) | Esophagus (C15) |
| Head and neck (C00-C14, C30-C32.9) | Accessory Sinuses (C31) |
| | Base of Tongue (C01) |
| | Larynx (C32) |
| | Hypopharynx (C13) |
| | Gum (C03) |
| | Floor of mouth (C04) |
| | Lip (C00) |
| | Lip, Oral Cavity & Pharynx, other & unspecified (C14) |
| | Major Salivary Glands, other & unspecified (C08) |
| | Mouth, other & unspecified (C06) |
| | Nasal Cavity & Middle Ear (C30) |
| | Nasopharynx (C11) |
| | Oropharynx (C10) |
| | Palate (C05) |
| | Parotid Gland (C07) |
| | Pyriform Sinus (C12) |

| | |
|---|---|
| | Tongue, other & unspecified (C02)<br>Tonsil (C09) |
| Hodgkin lymphoma (9650-9667) | Hodgkin Lymphoma (C) |
| Kidney and renal pelvis (C64.9-C65.9) | Kidney (C64)<br>Renal Pelvis (C65) |
| Leukemia (C91-C95, C90.1) | Leukemia (C) |
| Liver (C22.0, C22.2-C22.4, C22.7) | Biliary Tract, other & unspecified (C24)<br>Liver & Intrahepatic Bile Ducts (C22) |
| Lung and bronchus (C34) | Bronchus/Lung (C34) |
| Melanoma (C43) | Melanoma – other (C)<br>Melanoma of Eye (C69)<br>Melanoma of Skin (C44) |
| Multiple myeloma (C90.0, C90.2, C90.3) | Multiple Myeloma & Plasmacytoma (C) |
| Non-Hodgkin lymphoma (C82-C86) | Non-Hodgkin Lymphoma (C) |
| Ovary (C56) | Ovary (C56) |
| Pancreas (C25) | Pancreas (C25) |
| Prostate (C61) | Prostate Gland (C61) |
| Soft tissue including heart (C38.0, C47, C49) | – |
| Stomach (C16) | Stomach (C16) |
| Testis (C62) | Testis (C62) |
| Thyroid (C73) | – |
| Uterus (C54-C55) | Other Uterus & Uterus, NOS (C54-C55)<br>Endometrium (C54) |
| All other cancers (All sites C00-C80 not listed above, C97) | Anus & Anal Canal (C21)<br>Bones, Joints & Articular Cartilage of Limbs (C40)<br>Bones, Joints & Articular Cartilage of Other Sites (C41)<br>Immunoproliferative Diseases (C) |

| |
|---|
| Heart, Mediastinum & Pleura (C38) |
| Gallbladder (C23) |
| Female Genital Organs, other & unspecified (C57) |
| Digestive Organs, other & ill-defined (C26) |
| Male Genital Organs, other & unspecified (C63) |
| Meninges (C70) |
| Other Hematopoietic & Reticuloendothelial (C) |
| Penis (C60) |
| Placenta (C58) |
| Ureter (C66) |
| Urinary Organs, other & unspecified (C68) |
| Vagina (C52) |
| Vulva (C51) |

Abbreviations: ACR, Alberta Cancer Registry; CCS, Canadian Cancer Statistics.

**Supplemental Table 2.** Standardized incidence ratios (SIR) and 95% confidence intervals for risk of a different-site subsequent primary cancer (SPC) by age at first primary cancer and sex.

| | Females | | Males | |
|---|---|---|---|---|
| | **Survivors** | **SIR (95% CI)** | **Survivors** | **SIR (95% CI)** |
| **18-24** | 420 | 20.88 (11.07-33.78) | 613 | 4.84 (1.53-10.02) |
| **25-29** | 685 | 7.78 (4.52-11.91) | 635 | 11.52 (6.57-17.86) |
| **30-34** | 1,089 | 6.67 (4.67-9.03) | 851 | 4.71 (2.50-7.62) |
| **35-39** | 1,761 | 4.00 (3.03-5.10) | 1,024 | 5.55 (3.68-7.79) |
| **40-44** | 3,241 | 3.05 (2.56-3.59) | 1,678 | 5.20 (4.05-6.50) |
| **45-49** | 5,127 | 2.54 (2.24-2.85) | 3,405 | 3.34 (2.87-3.86) |
| **50-54** | 6,848 | 2.30 (2.09-2.51) | 6,112 | 2.92 (2.66-3.18) |
| **55-59** | 7,532 | 1.99 (1.84-2.16) | 9,451 | 1.99 (1.86-2.12) |
| **60-64** | 8,005 | 1.71 (1.59-1.84) | 10,637 | 1.78 (1.69-1.88) |
| **65-69** | 7,522 | 1.60 (1.49-1.71) | 11,169 | 1.39 (1.32-1.46) |
| **70-74** | 6,822 | 1.38 (1.29-1.49) | 9,399 | 1.34 (1.27-1.40) |
| **75-79** | 5,883 | 1.48 (1.37-1.59) | 7,948 | 1.22 (1.15-1.29) |
| **80-84** | 4,496 | 1.09 (0.98-1.21) | 5,180 | 1.20 (1.11-1.29) |
| **85-89** | 2,735 | 1.18 (1.02-1.36) | 2,484 | 1.15 (1.00-1.30) |
| **90+** | 1,174 | 1.19 (0.88-1.55) | 767 | 0.93 (0.66-1.24) |

Abbreviations: CI, confidence interval; SIR, standardized incidence ratio.

**Supplemental Table 3.** Standardized incidence ratios (SIR) and 95% confidence intervals for the risk of a different-site subsequent primary cancer (SPC) by income quartiles and urban/rural location at time of the FPC diagnosis.

| | Survivors | Obs / Exp | SIR (95% CI) |
|---|---|---|---|
| **Income quartiles** | | | |
| **Q1 (lowest)** | 34,073 | 3,447 / 2,146 | 1.61 (1.55-1.66) |
| **Q2** | 34,396 | 3,387 / 2,235 | 1.52 (1.46-1.57) |
| **Q3** | 32,998 | 3,276 / 2,064 | 1.59 (1.53-1.64) |
| **Q4 (highest)** | 33,111 | 3,155 / 2,101 | 1.50 (1.45-1.55) |
| **Residence at diagnosis** | | | |
| **Urban** | 110,879 | 10,868 / 6,981 | 1.56 (1.53-1.59) |
| **Rural** | 23,801 | 2,409 / 1,570 | 1.53 (1.47-1.60) |

Abbreviations: CI, confidence interval; Exp, expected number of SPC; Obs, observed number of SPC; Q, quartile; SIR, standardized incidence ratios.

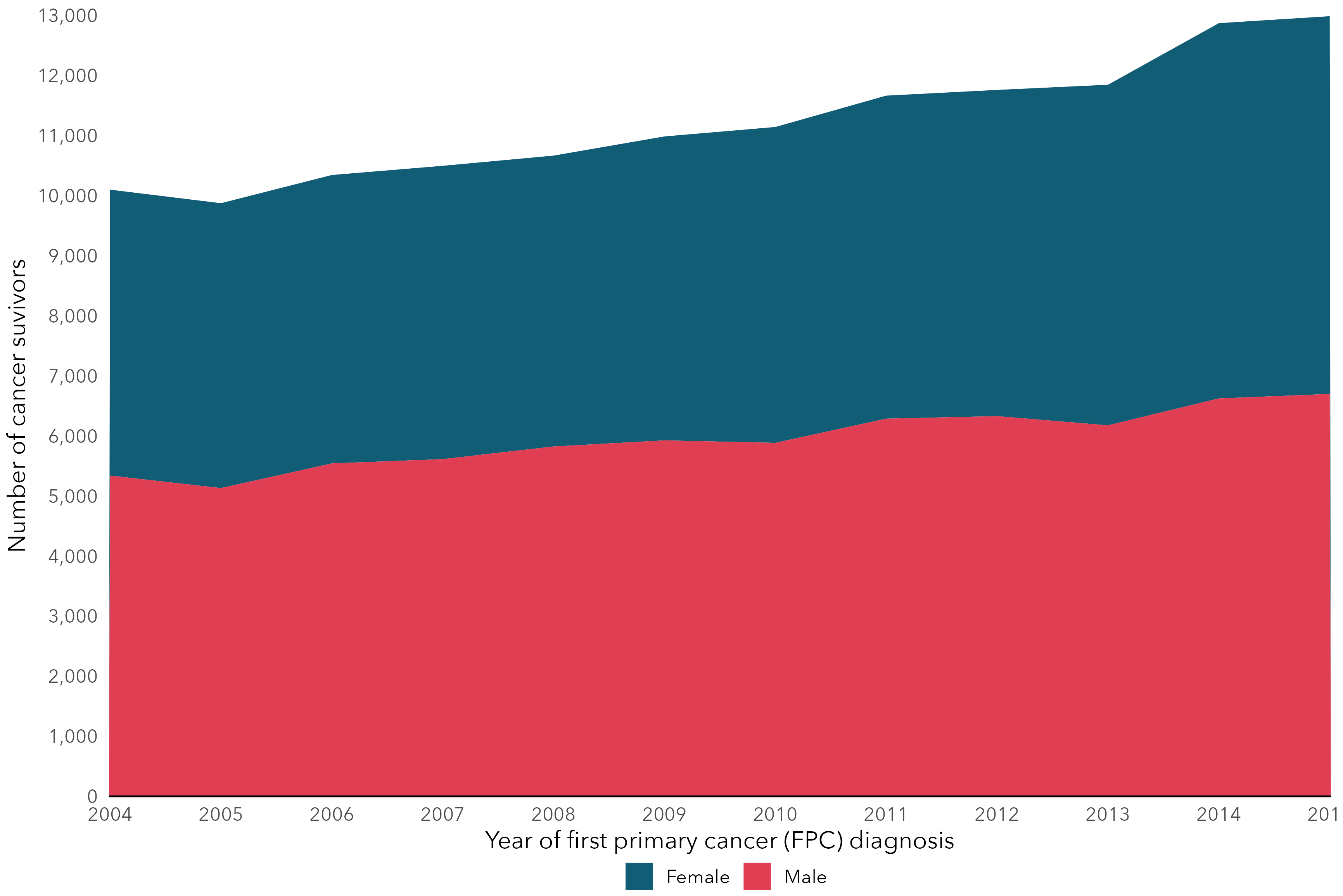


**Supplemental Figure 1.** The number of cancer survivors based on the year of the first primary cancer (FPC) diagnosis in Alberta, separately by sex.
